# Clinical Evaluation of a Multimodal On-Body Sensor Array

**DOI:** 10.64898/2026.07.29.26359254

**Authors:** Bright Nnadi, Sampath Rapuri, Carl Harris, John Rattray, Francesco Tenore, Charlene Gamaldo, Ralph Etienne-Cummings, Robert D. Stevens

## Abstract

Continuous, noninvasive blood pressure monitoring remains an unmet clinical need, particularly in the intensive care unit (ICU) where hemodynamically unstable patients need high-frequency monitoring. Invasive arterial catheterization represents the current standard of care for continuous blood pressure (BP) monitoring, but it carries risks and limits patient mobility. In this study, we evaluate the MOSAIC system, a novel multi-modal, multi-nodal wearable, wireless sensor system placed on multiple locations on the body, for continuous noninvasive BP estimation in a cohort of ICU patients. Unlike existing continuous BP sensors, the MOSAIC system offers an ideal form factor for continuous BP monitoring, enabling a fully untethered setup which minimally impacts activities of daily living. Leveraging sensor-derived biosignals to compute continuous BP, we determine the accuracy of our BP regression models using arterial line-derived blood pressure reading as a ground truth. Using a Light gradient boosted machine (LGBM)-based regression model, we demonstrate strong beat-to-beat agreement with a mean absolute error (MAE) of **5.66 *±* 5.94** mmHg for systolic BP (SBP) prediction and **2.45 *±* 2.87** mmHg for diastolic BP (DBP) prediction, and average ratio variability (ARV) of **0.527 *±* 0.185** and **0.489 *±* 0.170** for SBP and DBP, respectively, compared to linear and deep-learning regression baselines. Our findings demonstrate strong agreement between the predicted BP values and invasive, arterial-line BP measurements, supporting the feasibility of wearable, wireless, and cuffless blood pressure monitoring in high-acuity clinical settings.

## 1 Introduction

In clinical settings, BP is collected either through cuff-based, non-invasive sphygmomanometers that record intermittent, non-instantaneous measurements at intervals up to once every four hours[1], or through invasive continuous monitoring usually achieved through arterial catheter (“A-line”) insertion. Both approaches have significant limitations. Cuff-based BP measurements, while non-invasive in their acquisition, are not suitable for hemodynamically unstable patients in the intensive care unit (ICU) due to their low temporal frequency; for these patients, the standard of care typically requires continuous, high sampling rate blood pressure monitoring via the A-line [2]. In contrast, these invasive arterial catheter-based monitoring methodologies pose risks that include arterial thrombosis, infection, and hematoma; additionally, these devices tether the patient and require skilled placement and continuous oversight to ensure they remain appropriately placed and readings remain accurate [3–7].

Wearable BP sensors have emerged as a promising approach for continuous, non-invasive BP monitoring, with the potential to capture BP dynamics at high temporal resolution over extended timeframes without restricting routine activity. Nevertheless, current continuous BP technologies remain constrained by limitations in wearability, calibration burden, tethering, motion tolerance, clinical usability, and validation against invasive reference standards. Consequently, a noninvasive, untethered wearable BP monitoring solution that preserves patient mobility and integrates smoothly into clinical workflows remains an important unmet need, particularly in critically ill populations [8]. Here, we assess how well this untethered wearable BP monitoring system can achieve clinically acceptable agreement with invasive arterial catheter BP measurements in critically ill patients.

## 2 Results

Data were collected from 54 critically ill ICU patients across diverse demographics over a four-month enrollment period at the Johns Hopkins Hospital, a large tertiary referral center. Following quality screening a total of 34 recordings were excluded: 15 due to motion artifact, 6 due to insufficient data collection (*≤* 20 minutes), and 13 due to failed signal quality analysis discussed as detailed in the Methods. The 20-minute duration threshold was established empirically as a pragmatic criterion to ensure sufficient data for both subject-specific model calibration and a meaningful post-calibration evaluation window. A final cohort of 20 participants was retained for analysis, whose demographics and per-patient estimation results are summarized in Table 1.

**Table 1:**
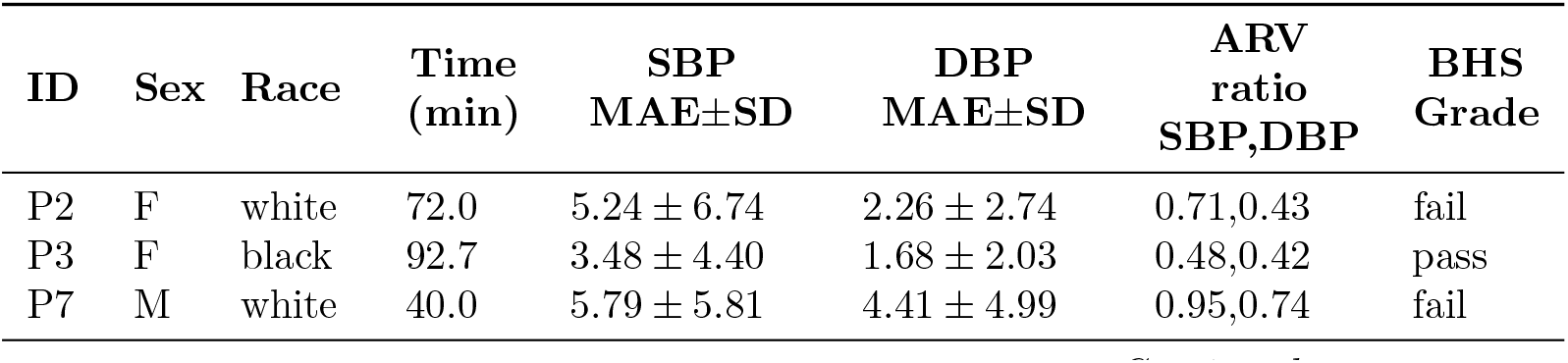
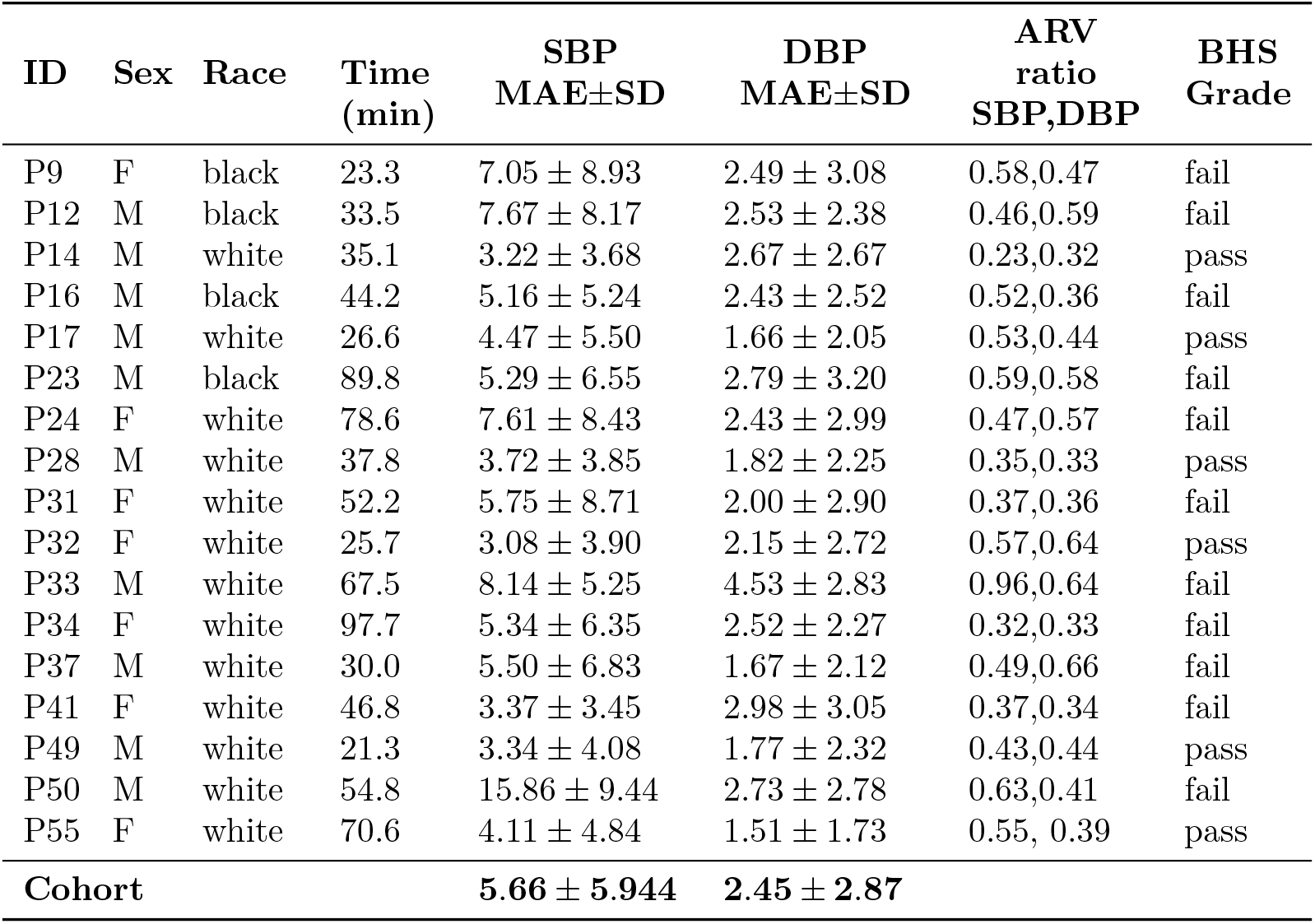
Participant demographics and estimated blood pressure analysis using the Light Gradient Boosting Machine (LGBM) model.

| ID | Sex | Race | Time<br>(min) | SBP<br>MAE±SD | DBP<br>MAE±SD | ARV<br>ratio<br>SBP,DBP | BHS<br>Grade |
| --- | --- | --- | --- | --- | --- | --- | --- |
| P2 | F | white | 72.0 | 5.24 ± 6.74 | 2.26 ± 2.74 | 0.71,0.43 | fail |
| P3 | F | black | 92.7 | 3.48 ± 4.40 | 1.68 ± 2.03 | 0.48,0.42 | pass |
| P7 | M | white | 40.0 | 5.79 ± 5.81 | 4.41 ± 4.99 | 0.95,0.74 | fail |

Table 1 – *continued*
| ID | Sex | Race | Time<br>(min) | SBP<br>MAE±SD | DBP<br>MAE±SD | ARV<br>ratio<br>SBP,DBP | BHS<br>Grade |
| --- | --- | --- | --- | --- | --- | --- | --- |
| P9 | F | black | 23.3 | 7.05 ± 8.93 | 2.49 ± 3.08 | 0.58,0.47 | fail |
| P12 | M | black | 33.5 | 7.67 ± 8.17 | 2.53 ± 2.38 | 0.46,0.59 | fail |
| P14 | M | white | 35.1 | 3.22 ± 3.68 | 2.67 ± 2.67 | 0.23,0.32 | pass |
| P16 | M | black | 44.2 | 5.16 ± 5.24 | 2.43 ± 2.52 | 0.52,0.36 | fail |
| P17 | M | white | 26.6 | 4.47 ± 5.50 | 1.66 ± 2.05 | 0.53,0.44 | pass |
| P23 | M | black | 89.8 | 5.29 ± 6.55 | 2.79 ± 3.20 | 0.59,0.58 | fail |
| P24 | F | white | 78.6 | 7.61 ± 8.43 | 2.43 ± 2.99 | 0.47,0.57 | fail |
| P28 | M | white | 37.8 | 3.72 ± 3.85 | 1.82 ± 2.25 | 0.35,0.33 | pass |
| P31 | F | white | 52.2 | 5.75 ± 8.71 | 2.00 ± 2.90 | 0.37,0.36 | fail |
| P32 | F | white | 25.7 | 3.08 ± 3.90 | 2.15 ± 2.72 | 0.57,0.64 | pass |
| P33 | M | white | 67.5 | 8.14 ± 5.25 | 4.53 ± 2.83 | 0.96,0.64 | fail |
| P34 | F | white | 97.7 | 5.34 ± 6.35 | 2.52 ± 2.27 | 0.32,0.33 | fail |
| P37 | M | white | 30.0 | 5.50 ± 6.83 | 1.67 ± 2.12 | 0.49,0.66 | fail |
| P41 | F | white | 46.8 | 3.37 ± 3.45 | 2.98 ± 3.05 | 0.37,0.34 | fail |
| P49 | M | white | 21.3 | 3.34 ± 4.08 | 1.77 ± 2.32 | 0.43,0.44 | pass |
| P50 | M | white | 54.8 | 15.86 ± 9.44 | 2.73 ± 2.78 | 0.63,0.41 | fail |
| P55 | F | white | 70.6 | 4.11 ± 4.84 | 1.51 ± 1.73 | 0.55, 0.39 | pass |
| **Cohort** |  |  |  | **5.66 ± 5.944** | **2.45 ± 2.87** |  |  |

### 2.1 Per-patient MAE tracking across models

Figure 1 summarizes the Mean Absolute Error (MAE) achieved by each model on a per-patient and cohort-wide basis.

**Fig. 1:**
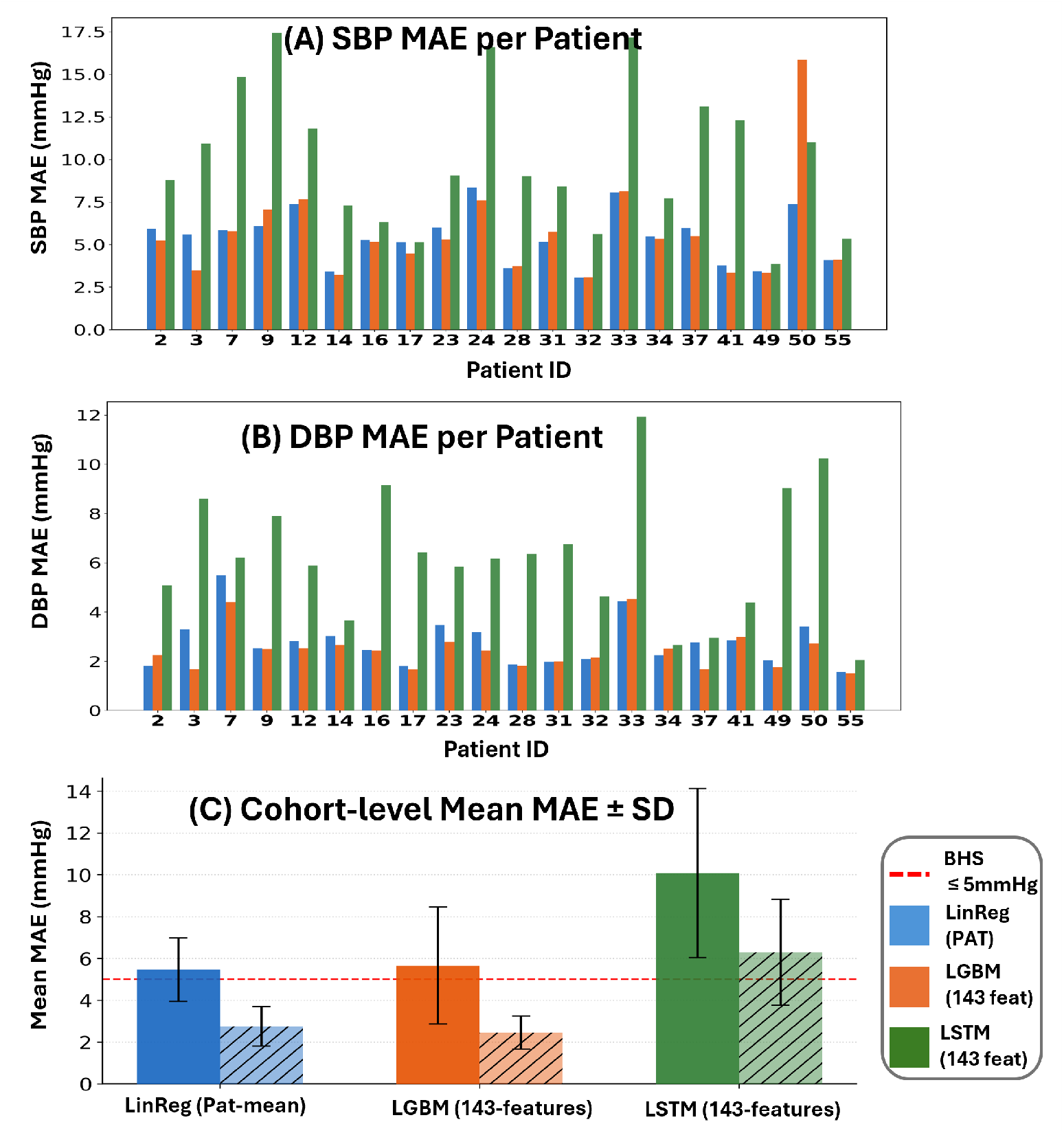
(A) Per-patient systolic blood pressure (SBP) mean absolute error (MAE); (B) diastolic blood pressure (DBP) mean absolute error (MAE) for all 20 participants across three models; (C) cohort-level mean of the mean absolute error (MAE) and *±* standard deviation (SD) for SBP (whole bar) and DBP (dashed bar). The dashed red line indicates the highest threshold (5%) adopted by the British Hypertension Society (BHS) for categorizing blood pressure monitors (cfr. [9]).

The **Linear Regression (PAT)** baseline, which uses only the mean pulse arrival time PAT extracted from the 15-second window preceding each BP measurement, achieves a cohort mean SBP MAE of 5.45 *±* 6.19 mmHg and DBP MAE of 2.75 *±* 3.10 mmHg. The **Light Gradient Boost Model (LGBM)** model, trained on the full 143-feature set derived from the synchronized ECG/PPG signals, yields comparable SBP accuracy (5.66 *±* 5.94 mmHg) with a marginal improvement in DBP (2.45 *±* 2.87 mmHg), remaining broadly within the British Hypertension Society (BHS) Grade A threshold of *≤* 5 mmHg for diastolic blood pressure [9]. The **Long Short Term Memory (LSTM)** model, a two-layer network with 128 hidden units, layer normalization, and dual regression heads (LSTM(143, 128, num layers=2, dropout=0.2)) produces substantially higher errors: SBP MAE of 10.09 *±* 8.47 mmHg and DBP MAE of 6.29 *±* 4.35 mmHg. This suggests that, for this dataset size, the sequence model overfits or fails to generalize beyond the training distribution despite regularization via dropout (*p*=0.2) and layer norm. Notable inter-patient variability is visible across all three models in Figure 1A–B, confirming that subject-specific physiology substantially influences prediction accuracy.

To characterize the structure of the 143-feature ECG/PPG representation, we performed PCA on the average within-subject feature correlation matrix. The feature space exhibited substantial low-dimensional structure: the first principal component explained 18.5% of variance and the first 15 components explained 66.0%. This suggests that the handcrafted feature set contains multiple partially redundant physiological dimensions rather than independent predictors, supporting the use of multivariate nonlinear models that can exploit covariance among timing, heart-rate, variability, and pulse-morphology-derived features.

To evaluate model performance under a realistic deployment scenario, a calibration strategy was applied: a short initial segment of each participant’s session (shaded gold in Figure 2 was used for subject-level adaptation, and the remaining signal constituted the unseen test period. All reported MAE and SD values are computed exclusively on the post-calibration test portion.

**Fig. 2:**
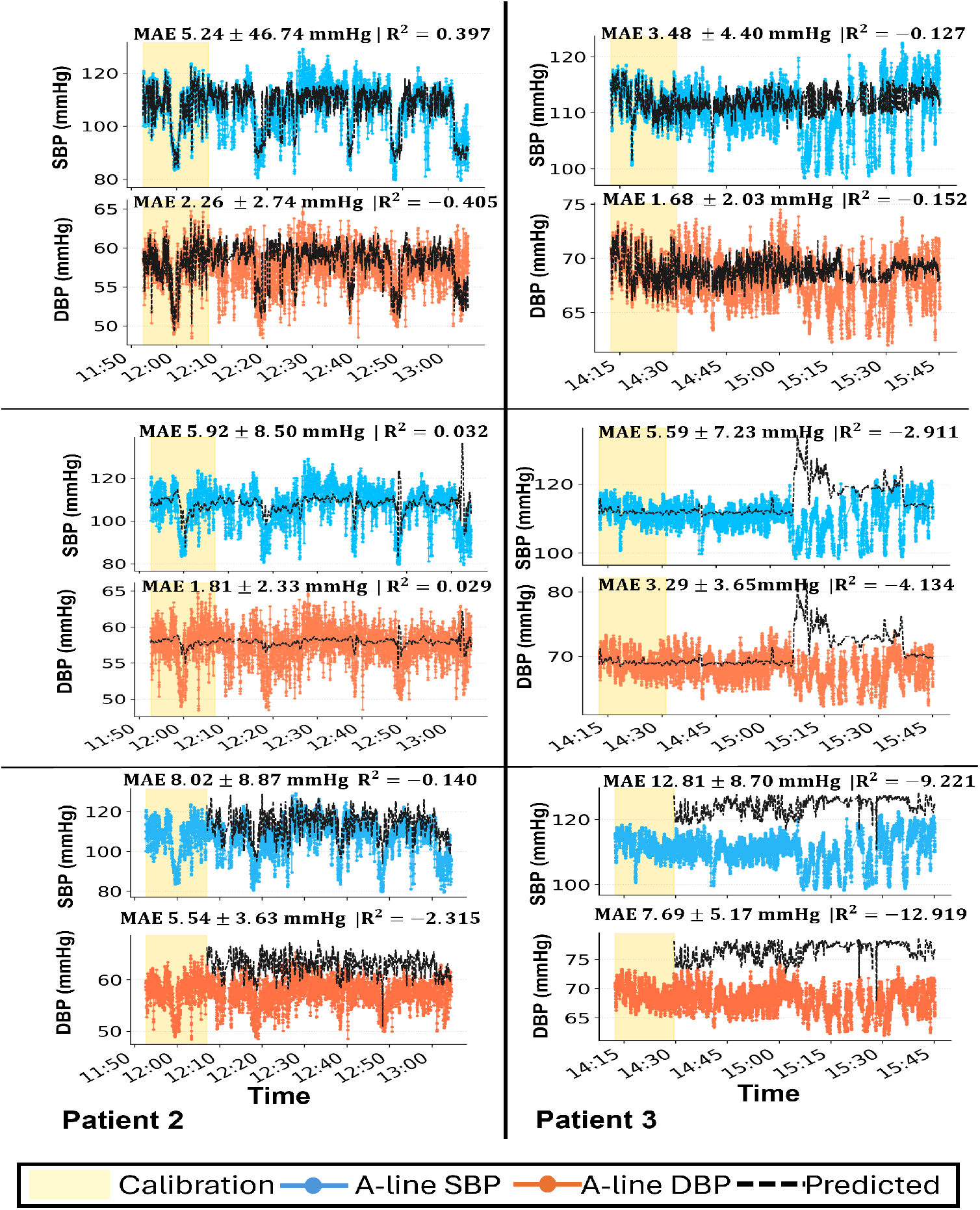
Continuous SBP and DBP estimation for two different patients (P2, P3) across three models: LGBM (top), Linear Regression PAT (middle), and LSTM (bottom). The gold-shaded region denotes the calibration period; metrics are evaluated on thepost-calibration test segment only.

Figure 2 illustrates continuous SBP and DBP estimation for two different patients (P2 and P3) across all three models. The LGBM model (Figure 2, top row) demonstrates the tightest coupling between predicted and arterial-line reference traces on both SBP and DBP for both patients, consistent with its lower per-patient MAE distribution observed in Figure 2. For P2, the LGBM achieves an SBP MAE of 5.24 mmHg and DBP MAE of 2.26 mmHg, while for P3 the corresponding values are 3.48 mmHg and 1.68 mmHg, respectively. The Linear Regression (PAT) model (middle row) captures the broad mean trend of the BP signal, with the predicted trace tracking the smoothed reference waveform throughout the evaluation window despite relying on a single feature. In contrast, the LSTM predictions (bottom row) exhibit a more smoothed trajectory that tends to regress toward the mean, particularly on SBP, which accounts for the substantially higher MAE values (P2: 8.02 mmHg SBP; P3: 12.81 mmHg SBP) and the large negative R^2^ values observed across both patients. Notable differences in recording duration and signal dynamics between the two patients further contextualise the per-patient results. P2 and P3 had session durations of 72.0 and 92.7 minutes, respectively, with the initial 20% of each session used for calibration. For P3, the latter half of the test period exhibits a marked shift in both BP range and beat-to-beat variability relative to the calibration window, a regime that all three models were not exposed to during training. This distributional shift is visibly reflected in the widening divergence between predicted and reference traces beyond the 15:10 mark in P3, where arterial-line SBP and DBP rise sharply while model predictions continue to track the lower baseline learned during calibration. This highlights if the calibration segment is drawn from a period of relative hemodynamic stability, the model may fail to generalize to the elevated variability and shifted BP ranges encountered later in the session, regardless of model complexity.

### 2.2 Average Real Variability (ARV) ratio analysis

While MAE quantifies absolute prediction accuracy, it does not capture whether a model faithfully reproduces the *dynamics* of beat-to-beat BP fluctuations. A model that predicts a constant mean value can achieve a low MAE if the patient’s BP is relatively stable, yet fail entirely to track physiologically meaningful variability. To address this, we compute the **Average Real Variability (ARV)** ratio, a metric derived from the ARV index originally proposed by Mena et al. as a clinically superior measure of short-term blood pressure variability compared to standard deviation [10, 11].

The ARV of a blood pressure sequence 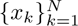 is defined as:

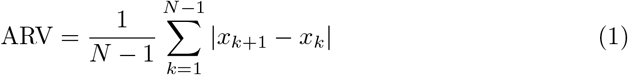

where *x_k_* denotes the blood pressure value at the *k*-th beat or time sample, *N* is the total number of blood pressure samples in the sequence, and *|x_k_*_+1_ *− x_k_|* is the absolute difference between two consecutive blood pressure measurements. A ratio of

1.0 indicates that the predicted sequence reproduces beat-to-beat variability identically to the reference; values approaching 0 indicate an over-smoothed prediction that tracks mean trends but suppresses physiological fluctuations. Figure 3 presents the ARV Ratio heatmap across all 20 patients for each model (SBP and DBP columns). **Model 1 (LGBM, 143 features)** achieves ARV ratios in the range 0.23–0.96 for SBP and 0.32–0.98 for DBP across the cohort, indicating that despite its strong MAE performance, the LGBM predictions retain a meaningful fraction of the true beat-to-beat dynamics for a number of patients, though considerable inter-patient variation is observed. **Model 2 (Linear Regression, PAT)** displays near-zero ARV ratios for the majority of patients (most values *≤* 0.28), revealing severe over-smoothing: the single-feature PAT-based predictor collapses toward a near-constant signal that does not track physiological BP variability, despite achieving competitive cohort-level MAE. This behavior is an inherent consequence of univariate linear-regression with only mean PAT as the input, the model has no access to beat-to-beat feature variation needed to reconstruct moment-to-moment BP fluctuations. This highlights a fundamental limitation of MAE as a sole evaluation metric for a model predicting a near-constant mean can score well on MAE while being clinically uninformative for continuous monitoring, underscoring the necessity of variability-preserving metrics such as the ARV ratio for evaluating beat-to-beat BP estimation systems. **Model 3 (LSTM, 143 features)** similarly suffers from low ARV ratios for most patients, with a few exceptions (notably P33 and P55, ARV ratio *≈* 0.96–1.18) where the model over-oscillates, suggesting prediction instability rather than genuine physiological tracking.

**Fig. 3:**
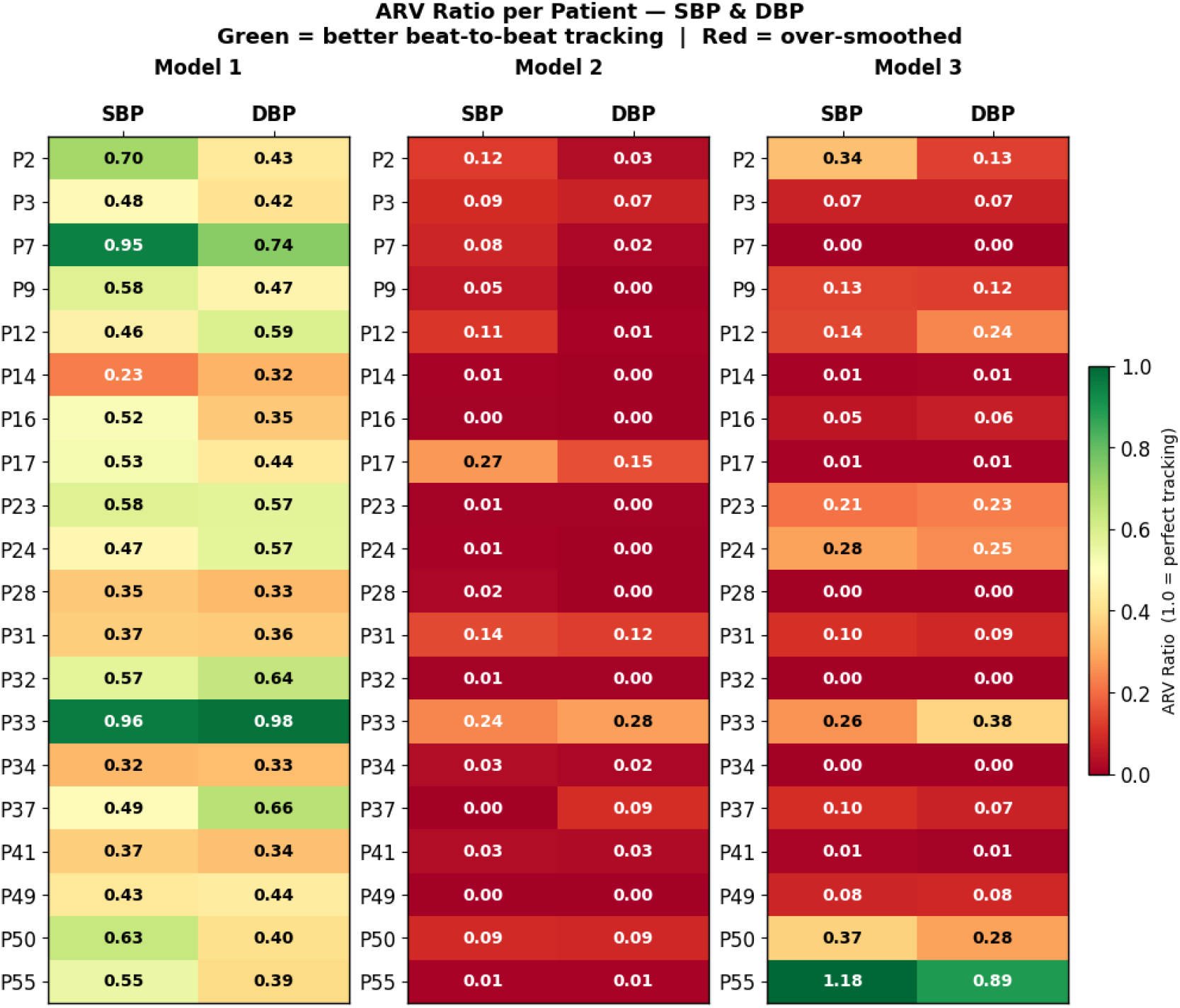
ARV Ratio heatmap for 20 participants across three models (SBP and DBP columns). Values close to 1.0 (green) indicate faithful beat-to-beat tracking; values near 0.0 (red) indicate over-smoothing.

### 2.3 statitical significance testing (Wilcoxon signed-rank test)

To illustrate how model comparisons manifest at the level of an individual recording beyond, the cohort-aggregated MAE reported in Table 1, we selected Patient P2 as a representative case to examine within-recording differences among the PAT, Light-GBM, and LSTM predictions. For this patient, per-beat absolute errors from each model were compared pairwise using a Wilcoxon signed-rank test, with Bonferroni correction applied across the three pairwise comparisons per outcome (SBP, DBP), yielding an adjusted significance threshold of *α*_Bonf_ = 0.0167. Results are summarized in Table 2.

**Table 2:**
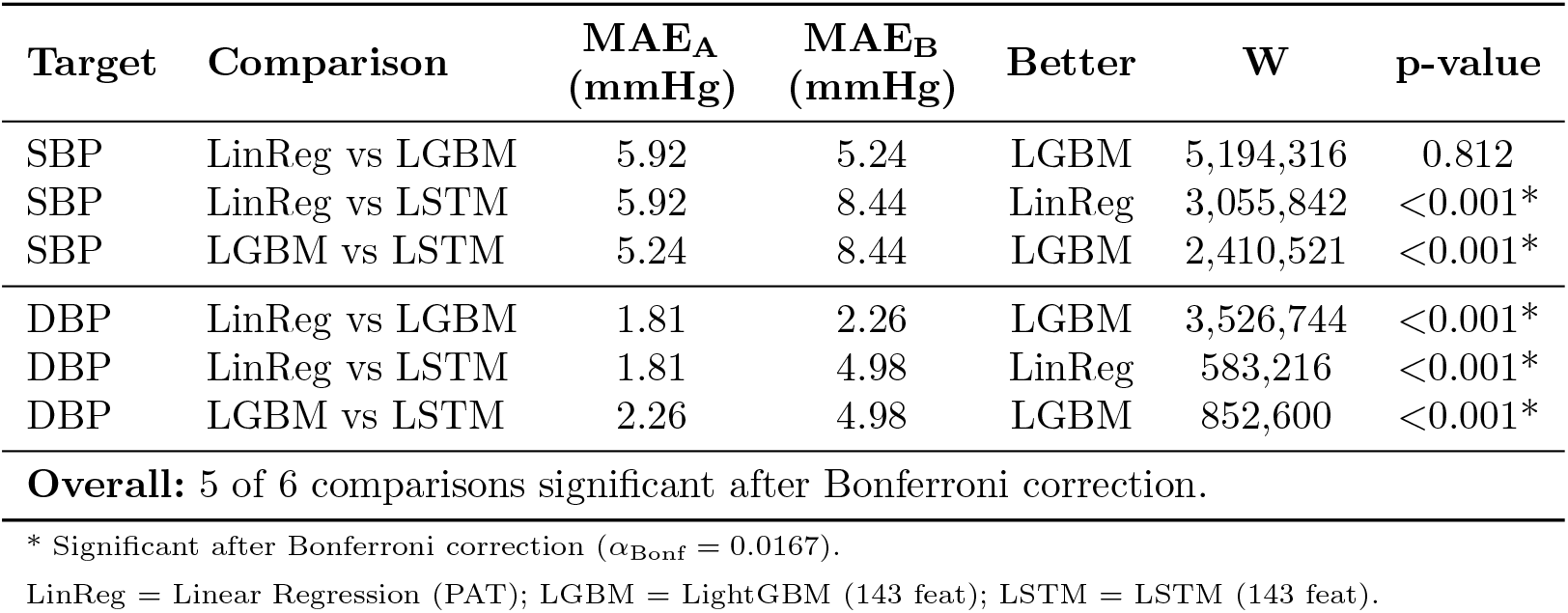
Pairwise Wilcoxon signed-rank test results with Bonferroni correction for Patient P2 (*α*_Bonf_ = 0.0167, 3 comparisons per outcome).

| Target | Comparison | MAE<sub>A</sub><br>(mmHg) | MAE<sub>B</sub><br>(mmHg) | Better | W | p-value |
| --- | --- | --- | --- | --- | --- | --- |
| SBP | LinReg vs LGBM | 5.92 | 5.24 | LGBM | 5,194,316 | 0.812 |
| SBP | LinReg vs LSTM | 5.92 | 8.44 | LinReg | 3,055,842 | <0.001* |
| SBP | LGBM vs LSTM | 5.24 | 8.44 | LGBM | 2,410,521 | <0.001* |
| DBP | LinReg vs LGBM | 1.81 | 2.26 | LGBM | 3,526,744 | <0.001* |
| DBP | LinReg vs LSTM | 1.81 | 4.98 | LinReg | 583,216 | <0.001* |
| DBP | LGBM vs LSTM | 2.26 | 4.98 | LGBM | 852,600 | <0.001* |
| **Overall:** 5 of 6 comparisons significant after Bonferroni correction. |  |  |  |  |  |  |

\* Significant after Bonferroni correction ( $\alpha_{\text{Bonf}} = 0.0167$ ).
LinReg = Linear Regression (PAT); LGBM = LightGBM (143 feat); LSTM = LSTM (143 feat).

For **SBP**, the LightGBM and Linear Regression models were statistically equivalent (*W* = 5,194,316, *p* = 0.812), with a clinically negligible MAE difference of 0.68 mmHg (5.24 vs. 5.92 mmHg). Both models significantly outperformed the LSTM (LinReg vs LSTM: *W* = 3,055,842, *p <* 0.001; LGBM vs LSTM: *W* = 2,410,521, *p <* 0.001), which returned a substantially higher SBP MAE of 8.44 mmHg.

For **DBP**, the Linear Regression model achieved the strongest performance (MAE = 1.81 mmHg), significantly outperforming both LGBM (*W* = 3,526,744, *p <* 0.001) and LSTM (*W* = 583,216, *p <* 0.001). The LGBM model also significantly outperformed LSTM for DBP (*W* = 852,600, *p <* 0.001), with MAEs of 2.26 and 4.98 mmHg respectively. These findings highlight that despite the LightGBM model’s 143-feature advantage, the single-feature PAT-based Linear Regression model achieves statistically equivalent or superior accuracy for this patient, underscoring the importance of patient-specific evaluation.

## 3 Methods

### 3.1 Experimental Setup

Patients were enrolled under an IRB-approved protocol (IRB00384821) at the Johns Hopkins Hospital. Eligible participants were adults aged *≥* 18 years admitted to the ICU with an arterial catheter for continuous BP monitoring. Participants were required to have no contraindications to sensor placement.

Two wearable sensors (see Fig 4 for device schematic as described by Rattray et al. [12]) were placed on each patient by a study team member. The ECG sensor was secured to the chest at the fourth intercostal space along the midclavicular line using two ECG electrodes; the placement site was wiped with an alcohol pad prior to attachment to ensure electrode adhesion and signal quality. The PPG sensor was secured to the index finger of the hand opposite the arterial line insertion using an adjustable velcro strap. Recording sessions lasted upwards of 4 hours, with a study team member present to observe signal quality and log any interrupting events such as nursing staff interactions, ambulatory behavior, or other occurrences that may have affected sensor recordings.

**Fig. 4:**
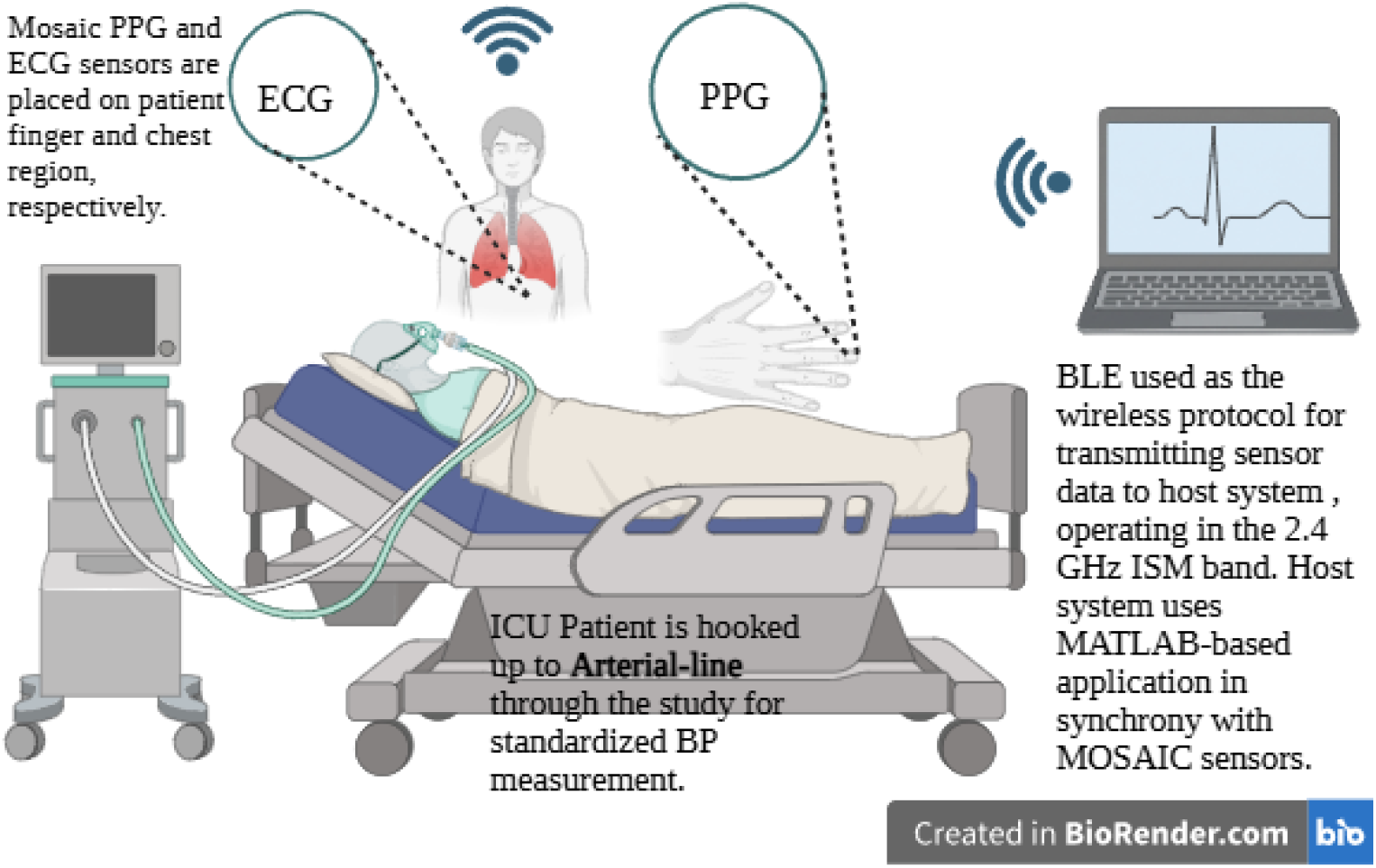
MOSAIC system overview for continuous, cuffless blood pressure estimation in ICU patients

During sensor recording periods, which are recorded at 250 Hz, all patients were bedridden and largely remained stationary, minimizing motion artifacts which are critical limitation given the known sensitivity of contact-PPG sensors to movement [13]. Ground truth ECG & PPG data were collected simultaneously at 250 Hz from 12-lead ECG leads and pulse oximetry sensors placed on each patient, respectively.

### 3.2 Signal Processing & Feature Extraction

Collected sensor data are preprocessed to improve signal quality. At the hardware level, there are analog filters that remove powerline interference and radio frequency artifacts [12]. Retrospectively, we further process raw ECG sensor data using a low-pass Butterworth filter with a cutoff frequency of 10 Hz in order to remove high-frequency noise. Likewise, PPG signals are low-pass filtered at 3 Hz.

Signal Quality Assessment was performed on both ECG and PPG signals to ensure data reliability and minimize artifacts that could confound subsequent analyses. The quality assessment employed a *template matching approach* implemented in the Neurokit2 Python Library. This method computes a continuous quality index by calculating the Pearson correlation coefficient between individual beat morphologies and a template representing the average beat morphology. The correlation coefficient *ρ* between each beat *s_r_*and template beat *s̄* was calculated as:

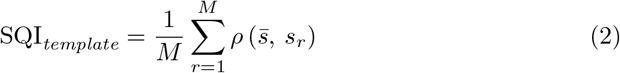

where *M* represents the total number of beats in the signal segment. Signal segments were evaluated using fixed-length windows of 10K samples (equivalent to a 40-second window at 250 Hz). For each window, the mean quality index was calculated by averaging the sample-level template-matching scores in equation 3. A quality threshold *τ ∈* [0, 1] was applied to the averaged window SQI, and a window was retained only if 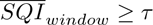.

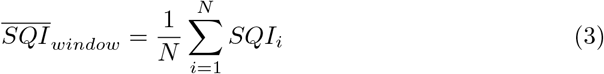

where *N* = 10, 000 samples and *SQI_i_* represents the quality index for the *i*-th sample.The window-based averaging approach reduces sensitivity to transient artifacts while maintaining adequate temporal resolution for identifying sustained periods of poor signal quality. Critically, temporal synchronization was enforced such that only PPG-ECG window pairs collected at identical timestamps were retained for analysis. This pairing strategy eliminated potential confounding from temporal misalignment and ensured that both physiological signals reflected the same cardiac cycles and hemodynamic events.

A related signal quality pipeline was applied in a concurrent MOSAIC study focused on continuous ABP waveform reconstruction in an overlapping ICU cohort [14]. In that work, wearable-to-reference ECG correlation was assessed over non-overlapping 5-minute blocks (rather than 40-second windows), and per-segment SQI thresholds of 0.6 were applied independently to ECG and PPG channels following R-peak-based temporal registration between sensor and bedside signals. Together, these two independently tuned SQA pipelines, applied to the same sensor platform but at different temporal granularities and for distinct downstream modeling tasks (beat-to-beat feature regression here vs. waveform reconstruction in [14]), reflect converging evidence that template-matching-based quality filtering is an effective strategy for excluding corrupted MOSAIC recordings prior to blood pressure modeling.

After the data were preprocessed, we extract manual features leveraging the mechanistic relationship between pulse arrival time (PAT) and blood pressure which has extensively been reported on in prior literature [4]. PAT is computed beat by beat by finding the interval between each detected R-peak from the filtered ECG using the Pan-Tompkins algorithm as implemented in NeuroKit2 [15] and the corresponding PPG pulse onset, which was computed as the maximum of the first derivative of the PPG waveform within a 200 ms search window surrounding each R-peak. The onset of each PPG pulse was defined as the point of maximum slope preceding the systolic peak. Beat pairs for which no valid PPG onset was identified within this window were not utilized for feature extraction.

Beyond PAT, we extracted additional morphological features from the PPG waveform as well as heart-rate variability (HRV) features from the ECG waveform (see Supplementary Table A1 for the full feature set). Features from the PPG waveform capture changes in arterial stiffness and compliance that influence BP dynamics [16], while the HRV features contain information about autonomic regulation of cardiac function which again influences BP [17, 18].

### 3.3 Modeling

We evaluate two categories of patient-specific models that build on the known PAT-based correlation with BP as well as incorporate other ECG and PPG-derived features to enrich the regression model. As a baseline, we employ a linear regression model using PAT as the sole predictor variable, motivated by the strengths of its easy interpretability and its strong correlation to our outcome BP variable.

From here, we incorporate additional features to enrich our analysis (see Supplementary Table A1) and create a multivariate linear model. To better capture nonlinearities in the enriched feature space and inter-feature interactions, we incorporate popular nonlinear regression models ranging from classical gradient boosting algorithms [19] to neural network approaches popular across related ECG and PPG-based regression tasks [20, 21].

Across all models, we split each patient’s data into causal training and validation 20% of dataset and testing 80% of datasets; meaning that across each dataset split, we conserve the ordering with respect to time of each split. The training dataset comes before the validation dataset which is before the test dataset in time. The 20% calibration window was selected empirically by sweeping the proportion of each session allocated to calibration (5%, 10%, 15%, and 20%) and evaluating downstream test performance. Cohort-level MAE was largely insensitive to calibration window size beyond 10%, whereas the ARV ratio, our metric of beat-to-beat variability preservation, improved consistently and most substantially at the 20% window (Figure 5). Because MAE alone can be minimized by an over-smoothed, near-constant prediction that fails to track true BP dynamics, as established in Section 2.2, we prioritized the calibration window that best preserved physiologically meaningful variability over the one that minimized MAE in isolation, and accordingly adopted a 20% calibration segment for all subsequent analyses. **Model 1 - Linear Regression Using Pulse Arrival Time (PAT mean)**: As a physiologically motivated baseline, we employed a patient-specific linear regression model using the mean PAT as the sole predictor variable, following the preliminary validation by Rattray [12]. PAT is defined as the time interval between the R-peak of the ECG and the foot onset of the corresponding PPG pulse shown in figure (6), and has been extensively reported to exhibit a negative correlation with BP due to its mechanistic dependence on arterial pulse wave velocity. Finnegan *et al.* [22] demonstrated that PAT exhibits strong negative correlations with BP *r < −*0.8 in controlled settings, while Escobar Restrepo *et al.* [23] reported moderate correlations in an ICU-specific cohort *r* = *−*0.50 for SBP; *r* = *−*0.42 for DBP, motivating its use as a clinically interpretable baseline.

**Fig. 5:**
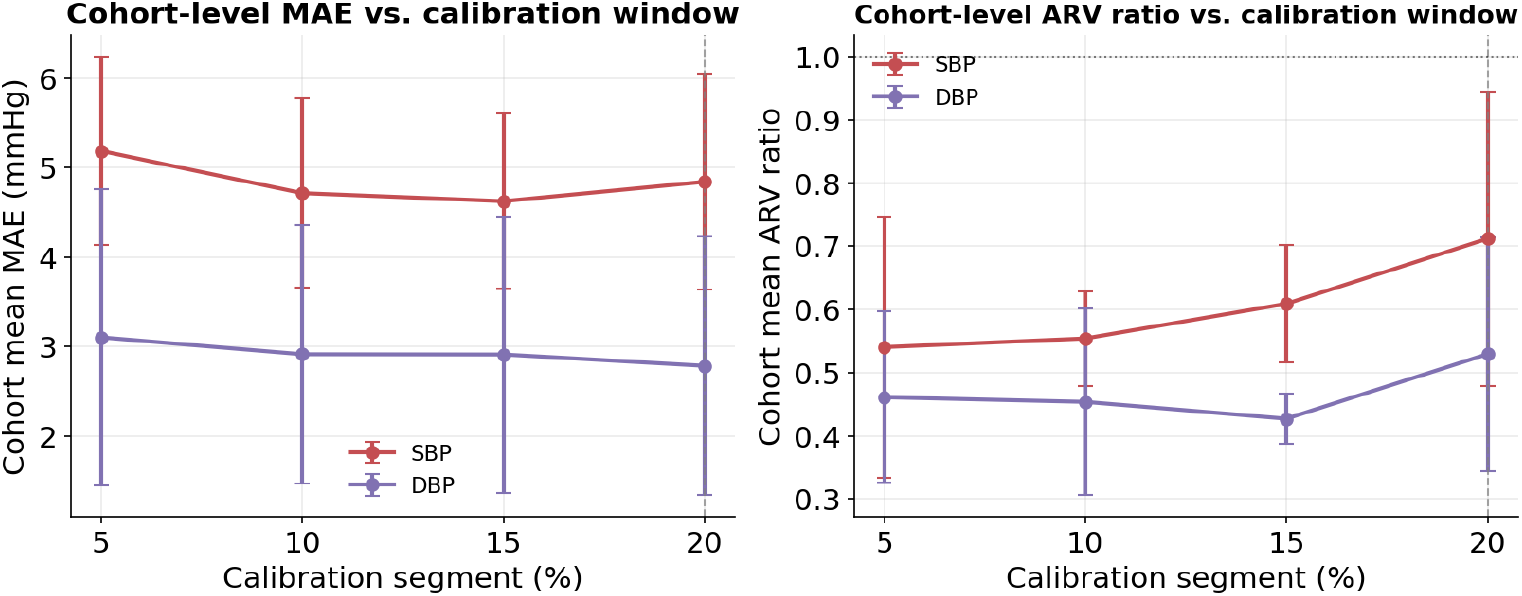
Cohort-level mean MAE (left) and ARV ratio (right) as a function of calibration segment size, shown with *±*1 SD error bars. Error stabilizes beyond a 10% calibration window, while ARV ratio continues to improve through 20%, motivating the calibration window size adopted in this study.

**Fig. 6:**
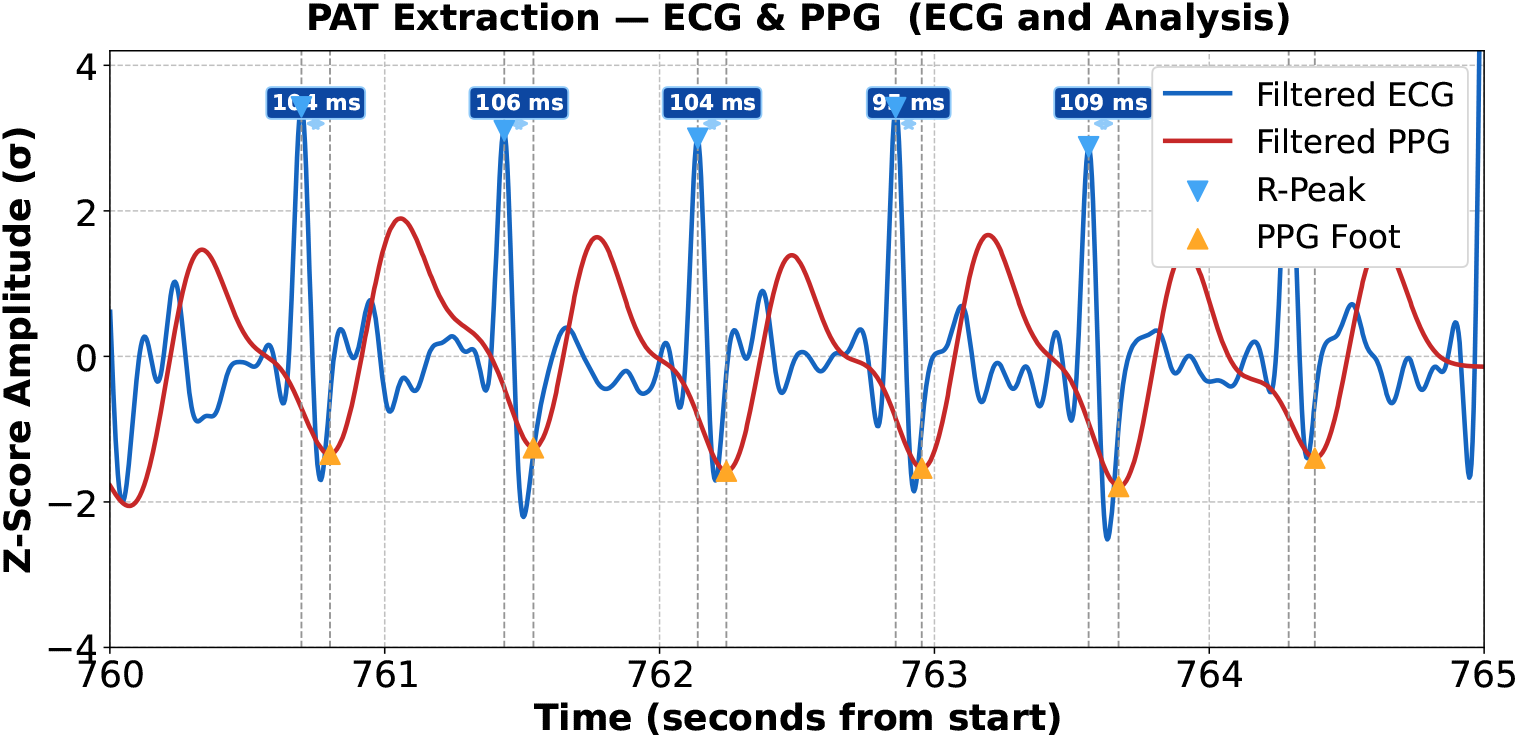
PAT extraction from synchronised ECG and PPG signals acquired by the MOSAIC wearable sensor.

**Model 2 - Non-Linear Regression Using LightGBM with Handcrafted Features**: To capture non-linear relationships between physiological features and blood pressure, we extended the feature set beyond PAT to include a comprehensive set of 143 handcrafted features derived from PPG morphology and heart rate variability (HRV). Specifically, beyond PAT, we extracted the timing intervals between the R-peak and the PPG systolic peak, PPG first derivative peak, and PPG second derivative peak, as well as intervals between successive PPG fiducial points displayed in figure (7). For each interval, 13 statistical aggregates were computed over a sliding beat window including mean, standard deviation, minimum, maximum, median, interquartile range, and recent change to encode both absolute timing and within-window temporal dynamics. HRV features including SDNN, RMSSD, NN50, pNN50, and heart rate slope were additionally incorporated to represent autonomic modulation of cardiac output, which has been shown to influence BP regulation.

**Fig. 7:**
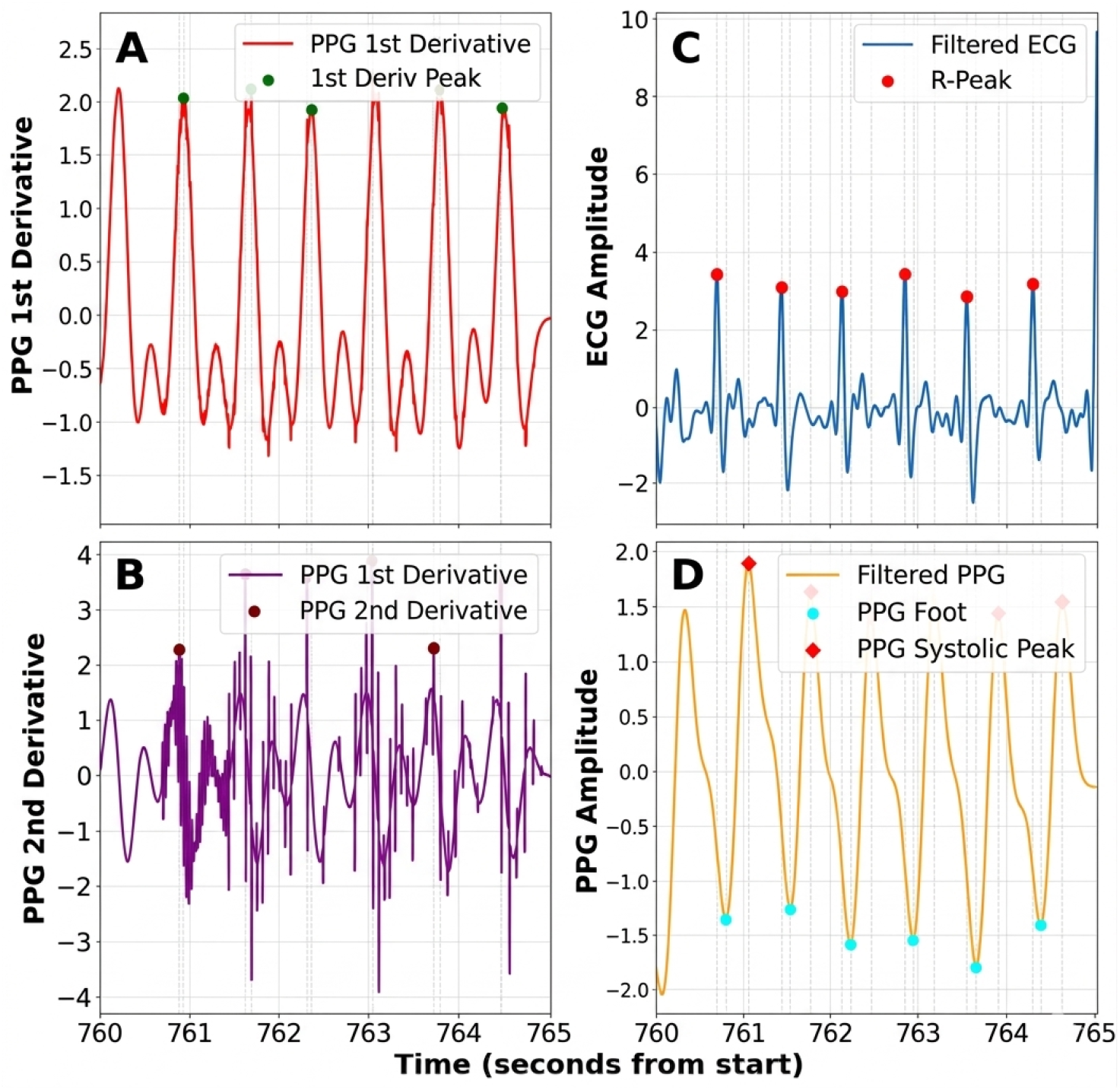
Morphological fiducial points extracted from the PPG waveform for non-linear feature construction

**Model 3 - Sequential Learning Using a Feature-Based LSTM** To exploit the temporal dynamics inherent in beat-to-beat physiological signals, we implemented a causal Long Short-Term Memory (LSTM) network operating over the same 143- feature representation used in Model 2. The LSTM receives a sliding window of 10 consecutive beat-level feature vectors as input and produces a one-step-ahead prediction of SBP and DBP, strictly preserving causality so that no future observations enter the input context. The architecture consists of a two-layer LSTM with a hidden size of 128, followed by LayerNorm and dropout regularization, and separate fully-connected regression heads for SBP and DBP. Each model was trained per patient using the Adam optimizer with a learning rate of 10*^−^*^3^, gradient clipping at norm 1.0, and early stopping with a patience of 15 epochs on the validation loss.

Finally, prior to training, a 5-sample moving average filter is applied to both the extracted beat-level features and the ground truth SBP and DBP waveforms to reduce sensitivity to transient noise.

## 4 Discussion

We demonstrate that continuous, noninvasive BP estimation is achievable using ECG and PPG signals acquired from the MOSAIC wearable sensor in a critically ill ICU population, extending our prior work in healthy volunteers [12] to a clinical cohort cohort characterized by high BP variability.

Among the three patient-specific regression models evaluated, the LightGBM model with 143 handcrafted features achieved the strongest overall performance, with an aggregate MAE of 5.66 *±* 5.94 mmHg for SBP and 2.45 *±* 2.87 mmHg for DBP predictions. Although the LightGBM MAE was comparable to the linear PAT baseline, the LightGBM model demonstrated stronger preservation of beat-to-beat blood pressure variability, with mean ARV ratios of 0.527 *±* 0.185 for SBP and 0.489 *±* 0.170 for DBP, compared with 0.066 *±* 0.079 and 0.046 *±* 0.071, respectively, for the linear PAT model. This indicates that, while both models produced similar absolute errors, the LightGBM model more faithfully tracked short-term BP dynamics, whereas the linear PAT approach tended to over-smooth the waveform and suppress physiologically relevant variability.

DBP predictions from both the linear and LightGBM models approach the AAMI/BHS clinical acceptability threshold of *≤* 5 mmHg MAE [24], with the Light-GBM model achieving a cohort DBP MAE of 2.45 *±* 2.87 mmHg and the Linear Regression model achieving 2.75 *±* 3.10 mmHg, as detailed in Table 1. SBP estimation proved more challenging, with both models returning cohort-level MAE values near the BHS threshold (LightGBM: 5.66 *±* 5.94 mmHg; Linear Regression: 5.45 *±* 6.19 mmHg), while the LSTM model exceeded the threshold substantially (SBP MAE: 10.09 *±* 8.47 mmHg; DBP MAE: 6.29 *±* 4.35 mmHg), consistent with the greater difficulty of estimating systolic peaks from morphological features alone.

To contextualise these errors, it is important to note that conventional sphygmomanometers which is the standard reference instrument in clinical practice are themselves not error-free. Under ideal laboratory conditions, aneroid and oscillometric devices have been shown to exhibit mean absolute measurement errors of up to 2.14 *±* 1.74 mmHg, with up to 18.6% of devices exceeding the *±*3 mmHg tolerance required by calibration standards [25, 26]. In critically ill patients specifically, oscillometric cuff measurements have been shown to underestimate arterial blood pressure by 6.7 *±* 9.7 mmHg on average relative to an invasive arterial line reference, with 26.4% of measurements deviating by *≥* 10 mmHg [27]. Furthermore, common procedural errors, including incorrect cuff sizing, improper arm positioning, and patient movement can introduce additional errors of 10–23 mmHg in routine clinical use [28, 29].

In this context, the DBP MAE of 2.45 *±* 2.87 mmHg and SBP MAE of 5.66 *±* 5.94 mmHg achieved by the LightGBM model (Table 1) are clinically competitive and in the case of DBP, below the inherent error floor of the conventional cuff, particularly under the challenging conditions of an ICU environment. These results support the feasibility of wearable, cuffless BP monitoring as a complement to invasive monitoring in the ICU, while also highlighting the remaining challenge of achieving consistent sub-5 mmHg SBP accuracy across all patients.

### 4.1 Comparison to Prior Work

Several cuffless BP monitoring approaches have been reported in the literature, differing substantially in sensor modality, form factor, and validation population. However, few have been validated against invasive arterial catheter ground truth in a clinical population where the the need for continuous BP monitoring is most acute but also where high BP variability poses a challenge for accurate prediction.

Ultrasound-based sensors have demonstrated strong BP estimation accuracy [30], but require tethered hardware that constrains patient mobility and limits practical continuous deployment in an ICU setting. Bioimpedance-based approaches offer an alternative noninvasive modality [31], though to our knowledge these have not been validated in ICU populations where high BP variability represents a challenge to model accurately. The Vitaliti neckband sensor has been evaluated in ICU patients [32], but its form factor, a rigid neckband, may be poorly tolerated for extended continuous wear and may interfere with clinical workflows.

In contrast to existing approaches, the MOSAIC sensor combines a chest ECG electrode and a finger-worn PPG sensor, both of which are unobstructive to daily living and minimally disruptive to regular clinical care. By validating this sensor against invasive arterial catheter ground truth in critically ill patients exhibiting high BP variability, we show that the MOSAIC sensor represents a favorable form factor for accurate, continuous BP estimation compared to previous sensor designs.

### 4.2 Limitations

While we present several strengths of the MOSAIC system, there are several limitations of this study. First, the MOSAIC sensor relies on contact PPG, which is known to be sensitive to skin tone and can introduce a systematic bias in our PPG-derived features that are used in the BP regression model [13]. As we did not stratify our patient cohort by skin tone, the generalizability of our findings across diverse skin tone remains unknown. All models evaluated in this study are patient-specific models, which means that a calibration period from each individual patient is required before the model can generate BP predictions. If deployed clinically, this approach would mean that each new patient would require an initial calibration period before reliable BP estimates can be generated, which may limit the MOSAIC system’s utility. Although we empirically selected a 20% calibration window based on ARV preservation, this represents a proportion of each recording rather than a fixed clinical duration. Future work should determine the minimum absolute calibration time required for reliable deployment.

## 5 Conclusion

We present a clinical evaluation of the MOSAIC wearable sensor system for continuous, noninvasive blood pressure estimation in critically ill ICU patients, demonstrating that patient-specific regression models trained on synchronised ECG and PPG signals can achieve beat-to-beat BP estimation competitive with the inherent error floor of conventional sphygmomanometers against an invasive arterial catheter ground truth. These findings extend the MOSAIC system’s prior validation in healthy volunteers [12] to a clinically challenging population characterized by high BP variability, and demonstrate that a chest ECG electrode and finger PPG sensor represent an unobtrusive, clinically viable form factor for continuous, cuffless BP monitoring. Future work should address the key limitations identified: the variable-length calibration period should be standardized to determine the minimum duration for clinically acceptable accuracy; the impact of skin tone on PPG-derived feature quality should be characterized; and cross-patient generalization via transfer learning or population-level models should be explored to remove the need for per-patient calibration before clinical deployment.

## Data Availability

MOSAIC sensor data is protected by Johns Hopkins University privacy restrictions, and sharing of these data will require a Data Use Agreement consented to by the University.

## Acknowledgements

This publication was made possible by the Johns Hopkins Institute for Clinical and Translational Research (ICTR), which is funded in part by Grant Number 1UM1TR004926 from the National Center for Advancing Translational Sciences (NCATS), a component of the National Institutes of Health (NIH), and NIH Roadmap for Medical Research. Its contents are solely the responsibility of the authors and do not necessarily represent the official view of the Johns Hopkins ICTR, NCATS or NIH.

This manuscript is the result of funding in whole or in part by the National Institutes of Health (NIH). It is subject to the NIH Public Access Policy. Through acceptance of this federal funding, the NIH has been given a right to make this manuscript publicly available in PubMed Central upon the Official Date of Publication, as defined by the NIH

This material is based upon work supported by the National Science Foundation Graduate Research Fellowship under Grant No. DGE2139757, awarded to CH. Any opinion, findings, and conclusions or recommendations expressed in this material are those of the authors(s) and do not necessarily reflect the views of the National Science Foundation.

## Appendix A Supplementary Tables

**Table A1:**
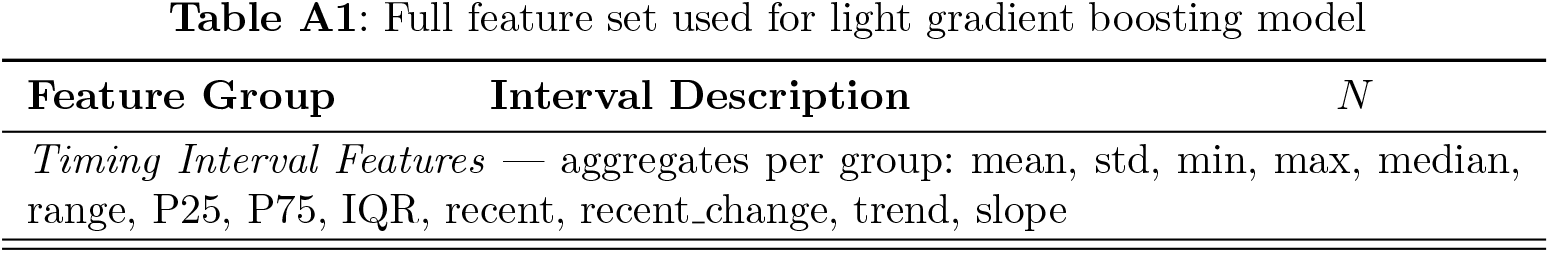
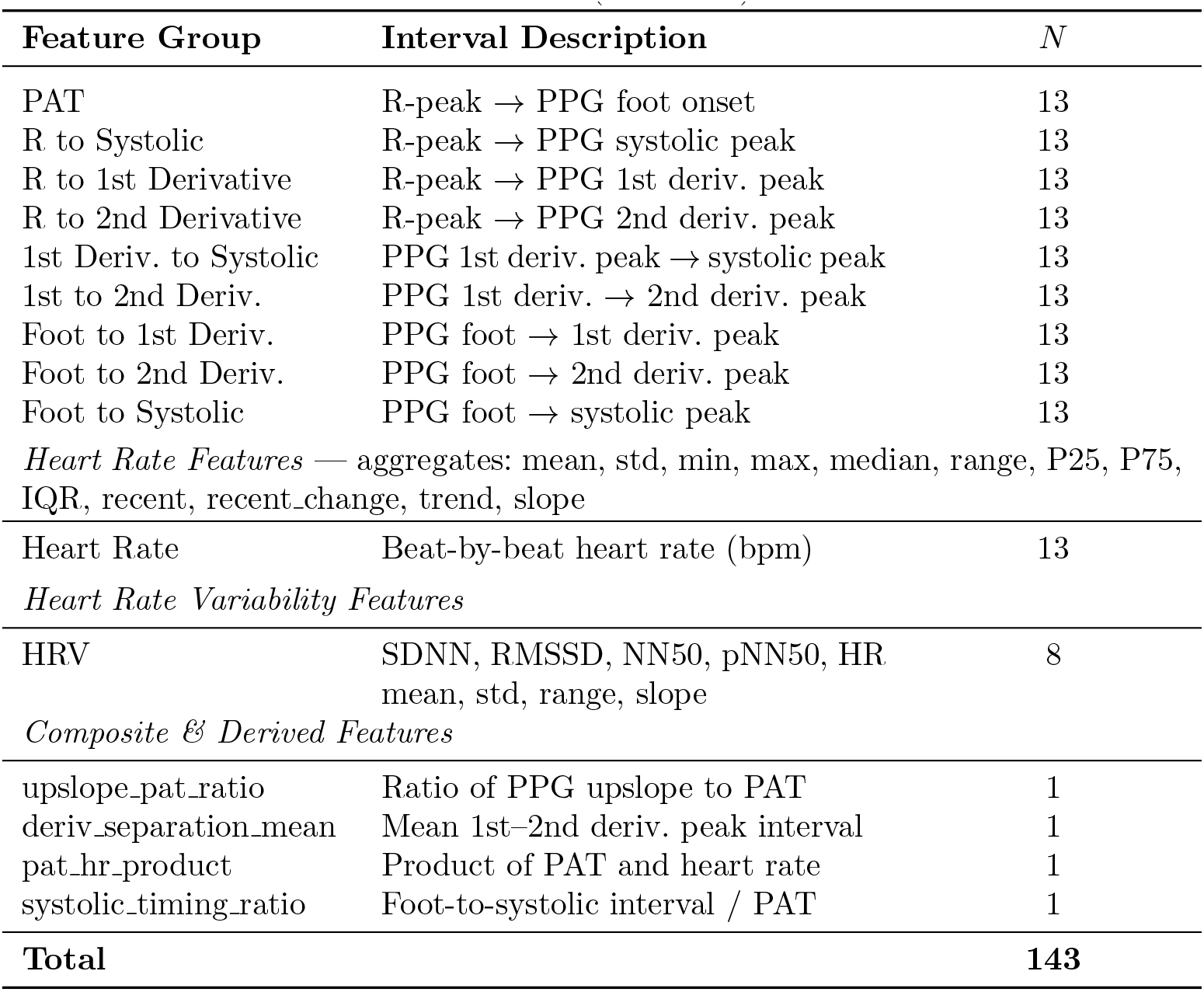
Full feature set used for light gradient boosting model.

| Feature Group | Interval Description | $N$ |
| --- | --- | --- |
| *Timing Interval Features* — aggregates per group: mean, std, min, max, median, range, P25, P75, IQR, recent, recent_change, trend, slope |  |  |

Table A1 (continued)
| Feature Group | Interval Description | $N$ |
| --- | --- | --- |
| PAT | R-peak → PPG foot onset | 13 |
| R to Systolic | R-peak → PPG systolic peak | 13 |
| R to 1st Derivative | R-peak → PPG 1st deriv. peak | 13 |
| R to 2nd Derivative | R-peak → PPG 2nd deriv. peak | 13 |
| 1st Deriv. to Systolic | PPG 1st deriv. peak → systolic peak | 13 |
| 1st to 2nd Deriv. | PPG 1st deriv. → 2nd deriv. peak | 13 |
| Foot to 1st Deriv. | PPG foot → 1st deriv. peak | 13 |
| Foot to 2nd Deriv. | PPG foot → 2nd deriv. peak | 13 |
| Foot to Systolic | PPG foot → systolic peak | 13 |
| *Heart Rate Features* — aggregates: mean, std, min, max, median, range, P25, P75, IQR, recent, recent_change, trend, slope |  |  |
| Heart Rate | Beat-by-beat heart rate (bpm) | 13 |
| *Heart Rate Variability Features* |  |  |
| HRV | SDNN, RMSSD, NN50, pNN50, HR<br>mean, std, range, slope | 8 |
| *Composite & Derived Features* |  |  |
| upslope_pat_ratio | Ratio of PPG upslope to PAT | 1 |
| deriv_separation_mean | Mean 1st–2nd deriv. peak interval | 1 |
| pat_hr_product | Product of PAT and heart rate | 1 |
| systolic_timing_ratio | Foot-to-systolic interval / PAT | 1 |
| **Total** |  | **143** |

**Fig. A1:**
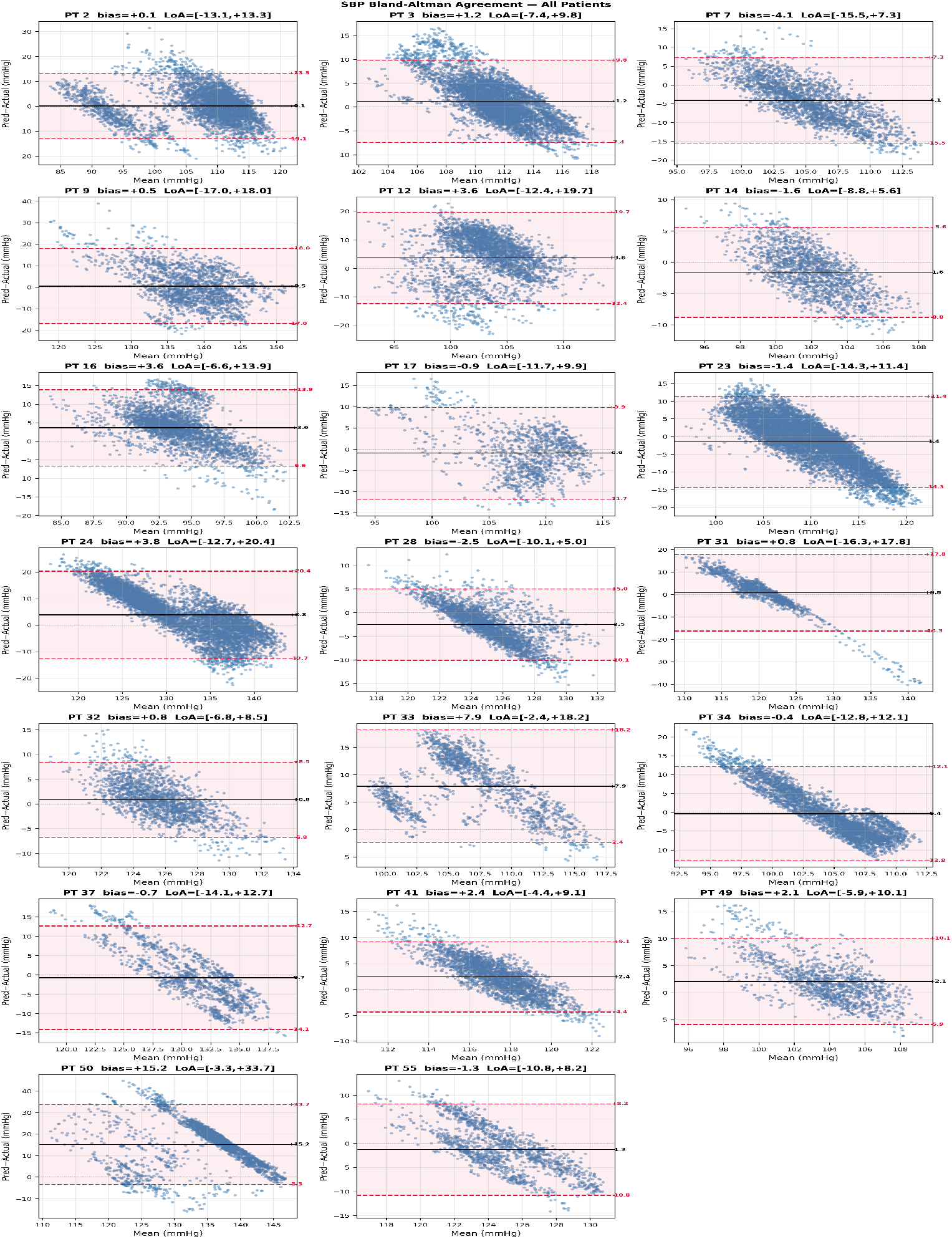
Bland-Altman plots of systolic blood pressure (SBP) estimation for all 20 patients. Each subplot shows the agreement between predicted and reference SBP values, where the x-axis represents the mean of predicted and actual measurements and the y-axis represents the signed difference (Predicted *−* Actual). The solid black line denotes the mean bias and the dashed red lines indicate the 95% limits of agreement (*±*1.96*σ*).

### A.1 Bland-Altman agreement analysis

We assessed model performance across each patient’s held-out causal test dataset, where the test data followed the training and validation data in time to prevent temporal data leakage. Mean absolute error (MAE) was the primary metric to assess the model’s fit to the data, using the arterial catheter-derived systolic and diastolic BP measurements as ground truth values.

In addition to per-patient MAEs, we performed a Bland–Altman analysis to quantify agreement between LightGBM-predicted and arterial-line BP measurements across all 20 participants

For each participant, the mean bias (average signed difference between predicted and reference BP) and 95% limits of agreement (LoA; mean bias *±*1.96 SD) were computed over the held-out test segment, and per-patient plots are shown in Figures A1 and A2.

For **SBP**, the cohort-level mean bias was +1.45 *±* 4.20 mmHg. Eighteen of 20 patients achieved a per-patient bias within *±*5 mmHg; the exception was PT 50, which exhibited a notably elevated bias of +15.2 mmHg and widest LoA of [*−*3.3, +33.7] mmHg, consistent with its high per-patient MAE of 15.86 *±* 9.44 mmHg reported in Table 1. Excluding PT 50, the cohort mean bias reduces to +0.73 *±* 2.75 mmHg. The mean 95% LoA spanned [*−*10.32, +13.23] mmHg across the cohort. For **DBP**, agreement was similarly strong, with a cohort mean bias of +0.27 *±* 1.68 mmHg and mean 95% LoA of [*−*4.98, +5.53] mmHg, again with all 20 patients within the *±*5 mmHg bias threshold. The presence of a mild proportional bias is visible as a negative slope in several Bland–Altman scatter plots that suggests that prediction accuracy decreases slightly at the extremes of the BP range, a common limitation of regression-based cuffless BP models in heterogeneous clinical populations [24, 25]

**Fig. A2:**
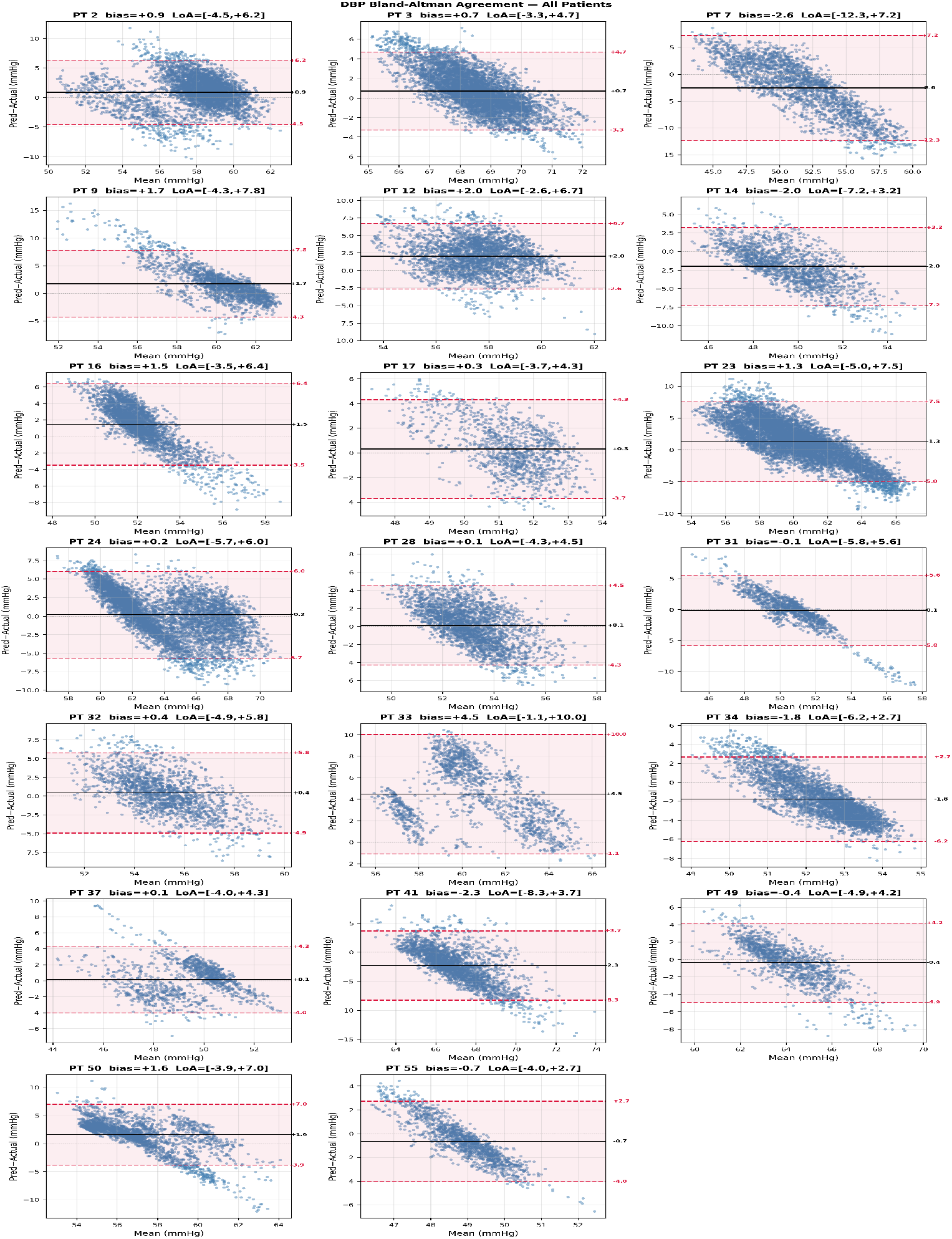
Bland-Altman plots of systolic blood pressure (DBP) estimation for all 20 patients. Each subplot shows the agreement between predicted and reference DBP values, where the x-axis represents the mean of predicted and actual measurements and the y-axis represents the signed difference (Predicted *−* Actual). The solid black line denotes the mean bias and the dashed red lines indicate the 95% limits of agreement (*±*1.96*σ*).

## Notes

### Competing Interest Statement

The authors have declared no competing interest.

### Author Declarations

The IRB of Johns Hopkins University gave ethical approval for this work under auspices of IRB00384821.

